# Household Socio-demographic Characteristics and Progress towards Attainment of Universal Health Coverage in Kilimanjaro, Tanzania

**DOI:** 10.1101/2022.06.18.22276172

**Authors:** Kanti Ambrose Kimario, Mikidadi Idd Muhanga, Kim Abel Kayunze

**Affiliations:** Department of Community Development and Gender, Moshi Co-operative University - Tanzania; Department of Development and Strategic Studies, College of Social Sciences and Humanities, Sokoine University of Agriculture -Tanzania

**Keywords:** Universal health coverage, socio-demographic characteristics, health services delivery quality, accessibility, affordability

## Abstract

Universal Health Coverage (UHC) attainment has been associated with households’ socio-demographic characteristics. Empirical findings have hardly dwelt on assessing the association between household socio-demographic characteristics and UHC attainment. This article assessed the association between socio-demographic features and attainment of UHC in Kilimanjaro Region, Tanzania. Specifically, the article: (i) analysed the perceptions on households’ socio-demographic characteristics in relation to UHC factors; (ii) determined the association between socio-demographic characteristics and UHC attainment; and (iii) estimated the level of UHC service coverage in the study area. The study employed a cross-sectional research design in which 384 households were selected through a multi-stage sampling approach and interviewed using a questionnaire. The Kruskal Wallis H Test and Mann Whitney U test were used as tests for association of socio-demographic variables and UHC factors. Geometric mean computation was used to estimate the level of UHC attainment. Results indicate; the level of UHC service coverage score of 69.9%, which is fairly good in relation to the WHO recommendation of 80%. Occupation (p = 0.012), general household health condition (GHHC) (p=0.039), health insurance membership (HIM) (p=0.039), and presence of non-communicable disease (p=0.032) were significantly associated with health services delivery quality. GHHC (p=0.041) was significantly associated with service accessibility. Income (p=0.000), occupation (p=0.000), education (p=0.004), health check-up frequency (p=0.001), and HIM (p=0.000) were significantly associated with health services affordability. Health services accessibility was not of great concern to most of the households. Some households could easily afford the cost of health services and others could not. Service providers, LGAs and MoH should promote affordability and accessibility of health services by the population regardless of their socio-demographic characteristics. Promotion of a single universal health insurance fund is essential for an improved progress to UHC attainment.

## 1. Introduction

Attainment of universal health coverage (UHC) entails sustainable access and quality delivery of healthcare services to all persons without being deprived financially [1, 2]. Continual realisation of UHC has been one of United Nations’ agenda for Sustainable Development Goals (SGDs), goal number 13, whereby nations and international institutions have prioritized UHC as one of the policy concerns [3]. UHC refers to an ongoing process involving improvement of people’s ability to access healthcare services in different contexts without experiencing financial hardship [4-6]. People’s capability and right to access healthcare services should not be deprived by their socio-demographic characteristics. Socio-demographic characteristics refer to individual and/or household features (social, demographic, and health-related) that place a person and/or a household in a situation to adequately or inadequately access healthcare services at a given time and environment. The characteristics referred to (in this study) include, among others, age, occupation, income, education level, and residence. Others are presence/absence of non-communicable disease, health check-up tendency, general household health condition, and health insurance membership. Since household’s socio-demographic features define the household’s capability and behaviour to access healthcare services, it is important to determine the association between the socio-demographic characteristics and the progress towards UHC attainment. The progress towards UHC attainment is defined by health services accessibility, affordability and services delivery quality.

Health services accessibility implies the situation where all those in need of health services can access the needed services without constraints in terms of geographical location, age, sex, economic status, education level or insurance membership [3]. This implies that all population groups are capable of accessing the needed health services within reasonable reach and affordable cost. Moreover, for the population in a given area to access the needed health services, there should be healthcare personnel, friendly infrastructure, equipment, and medicines available in the health facilities [7-9]. The government of Tanzania, in collaboration with other stakeholders in health (in the form of public-private partnership – PPP), for example, has helped in improving availability and accessibility of healthcare services. This is by reducing distance from healthcare seekers’ homes to the health facility, increased type of health services delivered to older people, and reduced medical fees, thus, making health services affordable [10-12].

Health services affordability involves reduction of healthcare costs sharing and fees while increasing pooled funds in terms of healthcare insurance for all. Health insurance for all is one of the mechanisms for making health services affordable to majority of the population in the formal and informal sectors. The presence of health insurance schemes which are friendly to all groups of people in the community makes health services affordable, and thus, more accessible. Several socio-demographic factors can be associated with individuals’ possibilities to access health insurance as well as pay for health services [13-15]. Community members’ ability to pay for health insurance, makes them assured of accessing health services with minimal or without limitation, based on individuals’ socio-demographic features. However, health services affordability is not itself enough without good health services delivery quality by healthcare providers.

Health services delivery quality is defined by the time/duration health facilities are open for services provision. Moreover, the duration that a patient can wait before being attended for service, and duration that a patient can be attended by healthcare personnel are indicators that can measure health services delivery quality. These indicators are closely linked with healthcare seekers’ services utilization. Health services utilisation (the extent to which healthcare seekers make use of health facilities for service consumption) is important in determining perceptions on health services delivery quality. Healthcare seekers’ socio-demographic factors such as education, residence, health insurance membership, age, and income, among others, do predict health services utilisation [15-18]. Socio-demographic features are important predictors of individual or household health because they define their capability to access health services.

Health is a foundation for human capital, a resource for social and economic development because healthy children can thrive in school and healthy adults can thrive in production [19]. Allocating resources for healthcare implies investing for human capital [6, 20]. The government of Tanzania, in collaboration with other stakeholders in the health sector, have been working hard to improve health services delivery quality to the community members at affordable cost and accessible environment. Some of the efforts, among others, include improving provision of primary healthcare which is easily accessible, affordable, sustainable, and gender considerate [21]. The efforts have been emphasized in the National Health Policy of 2017, the Health Sector Strategic Plan IV (HSSP IV) 2015-2020, the Tanzania Development Vision 2025, and the Five-Year Development Plan III (FYDP III) 2022-2026 [21, 22]. The efforts aim at increasing progress towards UHC.

As stated earlier, UHC has been associated with socio-demographic factors because they depict the status and capabilities of people to access healthcare services. Socio-demographic features are important predictors of individual or household health. Epidemiological profiles do change as fertility changes, incomes increase or decrease, populations age, and urbanization expands [23]. Moreover, non-communicable illnesses, accidents, and other external factors account for the increased burden of illness. Increased education, especially among women, can significantly be associated with health services use and health improvement [6, 15]. A study which involved 118 countries, found a strong relationship between UHC and structural factors (political stability, good governance, and socio-demographics) [6]. Besides, a study in Namibia found that socio-demographic features (education, sex, and wealth) were strongly associated with financial coverage and UHC attainment [24]. Moreover, studies in Ghana revealed that socio-demographic features (age, gender, education and population marginalisation, among others) were associated with financial and health services coverage [15, 25]. Several studies in Tanzania have assessed the influence and/association of social, economic and demographic characteristics on healthcare financing, access to specific healthcare services [26, 27], health insurance membership [28-30], among others. However, there is paucity of empirical evidence depicting the linkage between households’ socio-demographic characteristics and UHC attainment in Kilimanjaro. Thus, the study on which this article is based made an attempt to assess the association between socio-demographic factors and attainment of UHC in the study area.

## 2.0 Materials and Methods

### Study area

The study was carried out in four selected Districts/Councils of Kilimanjaro Region, namely, Rombo District Council (RDC), Moshi District Council (MDC), Hai District Council (HDC) and Moshi Municipal Council (MMC). Kilimanjaro Region is located in the North-Eastern part of Mainland Tanzania covering 13,209 square Kilometres. Administratively, the region has six district councils and one municipal council. The economic activities of the residents involve farming, animal keeping, tourism, and trade. Kilimanjaro has been a region with the highest human development index (0.75) in Tanzania based on human development report of 2017.

Kilimanjaro Region was purposefully selected for the study based on its highest score in the health system strength in Tanzania (with a z score of 3.8) [31]. This was measured in terms of healthcare infrastructure, health services utilization, health workers and quality. Considering this aspect, it was necessary to undertake the research in this area given that it also had health facilities under public-private partnership (PPP) operations as one of the measures for attaining UHC. It was important to know the contribution of PPP operations in the provision of health services in the region, which has been active since the 1990s. The four councils were also selected through purposive sampling, considering those with Council Designated or Voluntary Agency Hospitals operating under PPP service agreements with the LGAs.

### Research design

The study employed a cross-sectional research design through which data were collected at once from different sources in the selected councils. The design facilitates the collection of a body of quantitative and qualitative data about two or more variables (usually more than two), which are then examined to detect patterns of association [32, 33]. The design was chosen because it entails the collection of data on several cases at a time [32]. Moreover, the design was appropriate because the study intended to provide a snapshot of the linkages among households’ socio-demographic characteristics and UHC factors in the study area.

### Sample size and sampling procedure

The research sample size was 384 household respondents. Considering that the population size was large, Cochran’s formula [34, 35] was used to determine the sample size for the study. In determining the sample size, a z-value of 1.96, a p-value of 0.5, and a d-value of 5% (which is equivalent to 0.05) were used. This sample size is considered appropriate on the fact that “too large a sample implies a waste of resources, and too small a sample diminishes the utility of the results” [34-36].

### Sampling procedure

A multi-stage sampling technique [37, 38] was used involving selection of three clusters [14, 38], followed by proportionate random sampling of households at the district, ward, and village levels. Proportionate random sampling was used to obtain the number of households from each of the four selected councils. At the council level, five wards within health facility catchment areas were selected. In the selection, it was assumed that the household members in those wards were regular users of services in the selected health facilities. The number of households per ward “S1 Table **1** was also obtained proportionately as per the population and housing census of 2012 [39]. Out of 384 households, 86 were selected proportionately from RDC, 160 from MDC, 72 from HDC and 66 from MMC. At the ward level, proportionate sampling was also used to obtain the number of households. However, the criterion for selecting the wards was proximate distance (within a 10 km radius) from a hospital at Council level (PPP or government) because they were assumed to be the most frequent users of the hospital. At the village levels, simple random sampling technique was used to select households from a list of households in a village register. However, during data collection, if none of the household members had attended a health facility for any health-related issue in the previous 12 months (a criterion to select a household for interview), then the researcher would move to the next/nearby household. The reason is that such respondents could easily recall the nature of health services they received from a health facility.

Health facilities offering both inpatient and outpatient health services were considered. A total of 30 health facilities – Health Centres (HC), Council Hospitals (CH) and Council Designated Hospitals (CDH) were selected from the four councils. Out of the 30 health facilities, 20 were HCs and 10 were CHs and CDHs, distributed by ownership “S2 Table **2**. The PPP health facilities (involving two HCs and eight CDH/voluntary agency hospitals) were selected based on the presence of a signed PPP service agreement. The government health facilities involved 18 HCs and two hospitals at the district level. Dispensaries and clinics were excluded because the range and type of health services offered could not suffice the purpose of this study.

### Measurement of variables

The dependent variable for this study was UHC, and the independent variables were the households’ socio-demographic characteristics. Measurement of the dependent variable was customized from various authors [40-44], who assessed services access in terms of availability, affordability, accessibility, accommodation and acceptability. However, this paper considered three components of UHC which are health services accessibility, affordability and health services delivery quality. The three components were measured based on 5, 4, and 9 statements respectively, customized from other studies [40, 41, 43-45]. For those statements, the respondents were required to respond strongly disagree (1 point) …to… strongly agree (5 points). For health services accessibility, affordability, and health services delivery quality “S4 Table **4**, index summated scales made up of 5, 4, and 9 statements, respectively, were used to obtain median scores of the perceived health services accessibility, affordability, and service delivery quality. The scores were later used as inputs in the Kruskal Wallis H Test and Mann Whitney U test. Thus, it was possible to determine whether there were significant differences in UHC levels (the three constructs – health services accessibility, affordability and services delivery quality) between different categories of socio-demographic characteristics of the respondents. Thus, the dependent variable (and its constructs) was measured at scale level before being categorised into groups of ordinal measure. However, the independent variables (socio-demographic factors) were measured at the nominal and ordinal levels “S3 Table **3**.

### Data collection and analysis

For data collection see of the UHC coverage index, derived from geometric means (GM) of the tracer items grouped into four domains. Geometric mean was preferred in this context because of its ability to tolerate extreme values [23].

**Table 1**.

**Table 1:** Data collection and analysis matrix.

| Study objective | Data Collection | Data Analysis |
| --- | --- | --- |
| 1. To analyse household perceptions on socio-demographic characteristics of household respondents and UHC factors in the study area | **Method:** Household-based survey<br>**Tool:** Household-based survey questionnaire | Cross-tabulation to describe the perceptions of household's socio-demographic characteristics in relation to factors for UHC attainment in terms of frequencies and percentages.<br>**Analysis software:** IBM SPSS V. 23 |
| 2. To determine the association between various socio-demographic variables and UHC factors | **Method:** Household-based survey<br>**Tool:** Household-based survey questionnaire | **Kruskal-Wallis H test** (involving use of **Dunn's post hoc test** with the **Bonferroni correction** for pairwise comparison of the predictor variables with statistically significant difference)<br>**Mann-Whitney U test** was used to determine association between dichotomous predictor variables and UHC factors<br>**Analysis software:** IBM SPSS V. 23 |
| 3. To determine the extent/level of UHC in the study area | **Method:** Health facilities-based Survey<br>**Tool:** Health facilities-based questionnaire. Adapted from Tanzania's health facilities registry questionnaire and the WHO's Service availability and readiness assessment tool (SARA)[48, 49]. | Computation of UHC service coverage index derived from **Geometric Mean** of the tracer items and expressed in percentage.<br>**Analysis Software:** Microsoft Excel |

The socio-demographic characteristics (predictor variables) involved were age – categorised in age groups [46], income (categorised based on quartiles and mean as a cut-off point), household size, and education level. Another feature is health check-up frequency (categorised based on the WHOs recommended number of health check-up visits per person per year, considering 5 visits as a threshold). Other features are residence, occupation, health insurance membership, and presence of non-communicable diseases/illnesses, and general household health condition (GHHC). The GHHC was categorised into three categories: fair, good, and very good. Fair (if a household had a patient suffering from an illness at the time of household visit). Good (if a household had no patient at the time of household visit but have had one who has recovered from an illness in the past three months). Very good (if a household has no any patient and has not suffered from any illness in the past three months). To describe the predictor variables in relation to UHC factors, and using median as a cut-off point, the 25^th^, 50^th^, and 75^th^ quartiles were obtained. The quartiles were used to group each factor of the dependent variable into three levels Table 3:

#### Socio-demographic characteristics and UHC factors crosstabulation (n = 384)

The second objective involved determining the association between socio-demographic variables and attainment of UHC in the study area. Through SPSS, the Kruskal-Wallis H-Test (an alternative for ANOVA in parametric analysis) was used to determine differences in UHC levels between different categories of household’s socio-demographic characteristics. Moreover, Dunn’s post hoc test was done with the Bonferroni correction for the predictor variables found with statistically significant difference. The Kruskal-Wallis H-test was chosen because One-Way ANOVA was inappropriate for this study due to violation of two assumptions: the assumptions of normality of data (using Shapiro-Wilk Test) and the assumption of homogeneity of data/equal variance (using Levene’s Test). A Mann Whitney U test was also used to show the differences between dichotomous socio-demographic characteristics and UHC factors.

The third objective involved determining the level of UHC service coverage in the study area. To achieve this, an approach for tracking progress towards UHC attainment, as adopted by [23] and [1], from [47] was applied with some adjustments. The approach as in (Fig.1. **A Framework for calculating UHC service coverage index [**47]) involved computation of the UHC coverage index, derived from geometric means (GM) of the tracer items grouped into four domains. Geometric mean was preferred in this context because of its ability to tolerate extreme values [23].

**Fig. 1.**
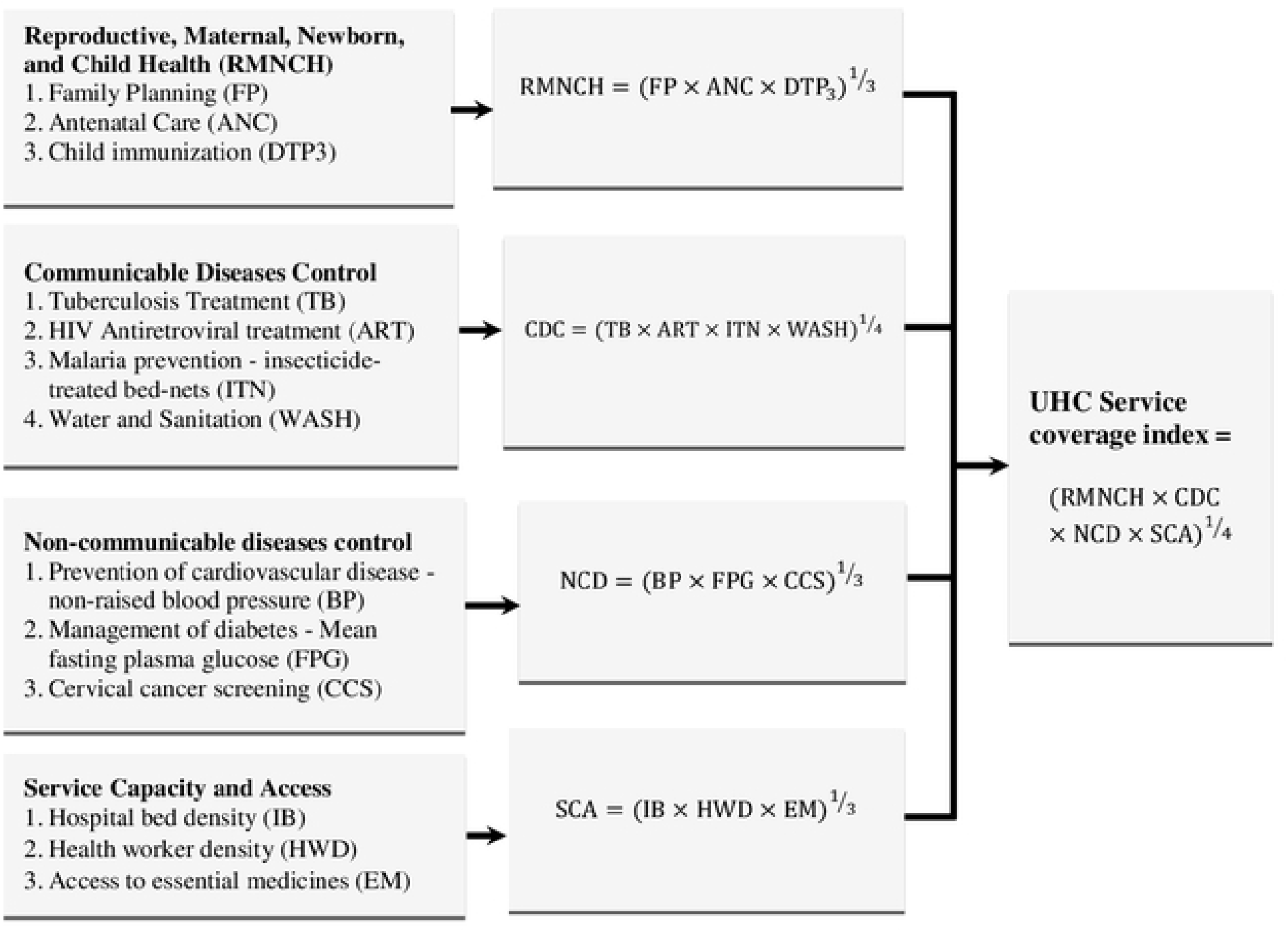
A Framework for calculating UHC service coverage index [47].

### Ethical approval and participation consent

A research clearance permit for this research was first obtained from the office of the Vice Chancellor of the Sokoine University of Agriculture with Ref. number SUA/ADM/R.1/8/589. Then a research permit was sought from the President’s Office Regional Administration and Local Government (PO-RALG) of Tanzania under the Regional Administrative Secretary office. The RAS issued a permit with ref. number FA.228/276/03/14 to be submitted to the District Administrative Secretary office in the four selected district councils who also issued a permit to conduct the research in the selected areas in each district council. At the household levels, an oral consent was sought from the household respondents before interview and were free to stop any time at their convenience. Ethical clearance was also sought from the District Medical Officer for data collection in the selected health facilities.

## 3.0 Results and discussion

### Households’ socio-demographic characteristics

The households’ socio-demographic characteristics were a combination of social (education level, income, economic activity), demographic (age, household size), and health-related (health insurance membership, presence of non-communicable disease, and general household health condition) characteristics. Detailed description of these socio-demographic characteristics is presented in Table 2.

**Table 2:**
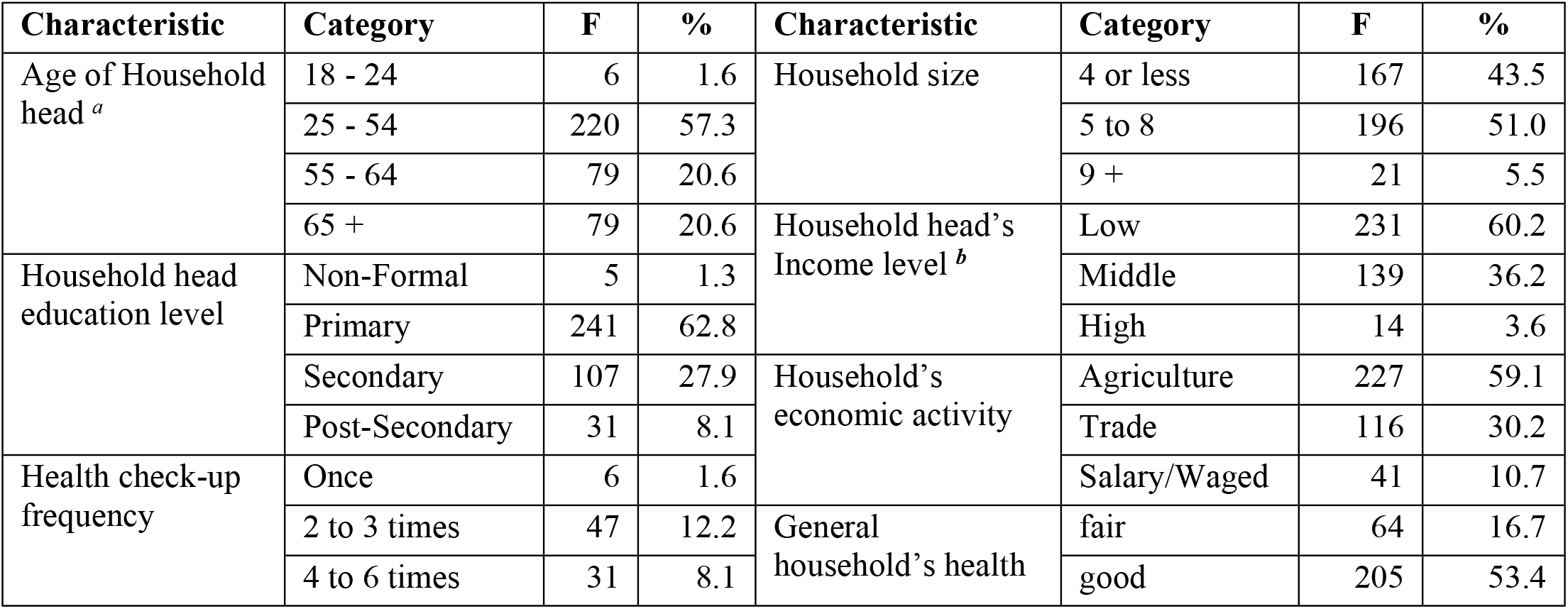
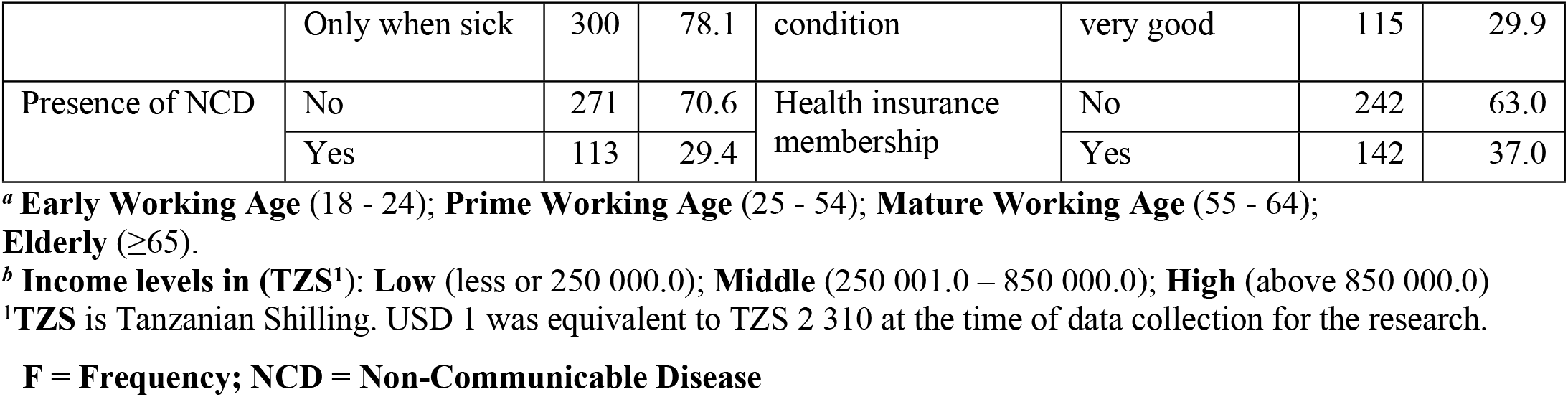
Households’ Socio-demographic characteristics (n = 384)

### Respondents’ Perceptions on household socio-demographic characteristics and UHC factors

Results in Table 3 indicate that health services accessibility was high (≥ 88%) almost among all household respondents distributed in different socio-demographic characteristics. This indicates a positive perception on health services accessibility among the household respondents. The implication is that health services were accessible in terms of geographical location, infrastructure friendliness, among others. Thus, based on these perceptions, healthcare services accessibility was not of great concern to majority of the household respondents in the study area.

**Table 3:**
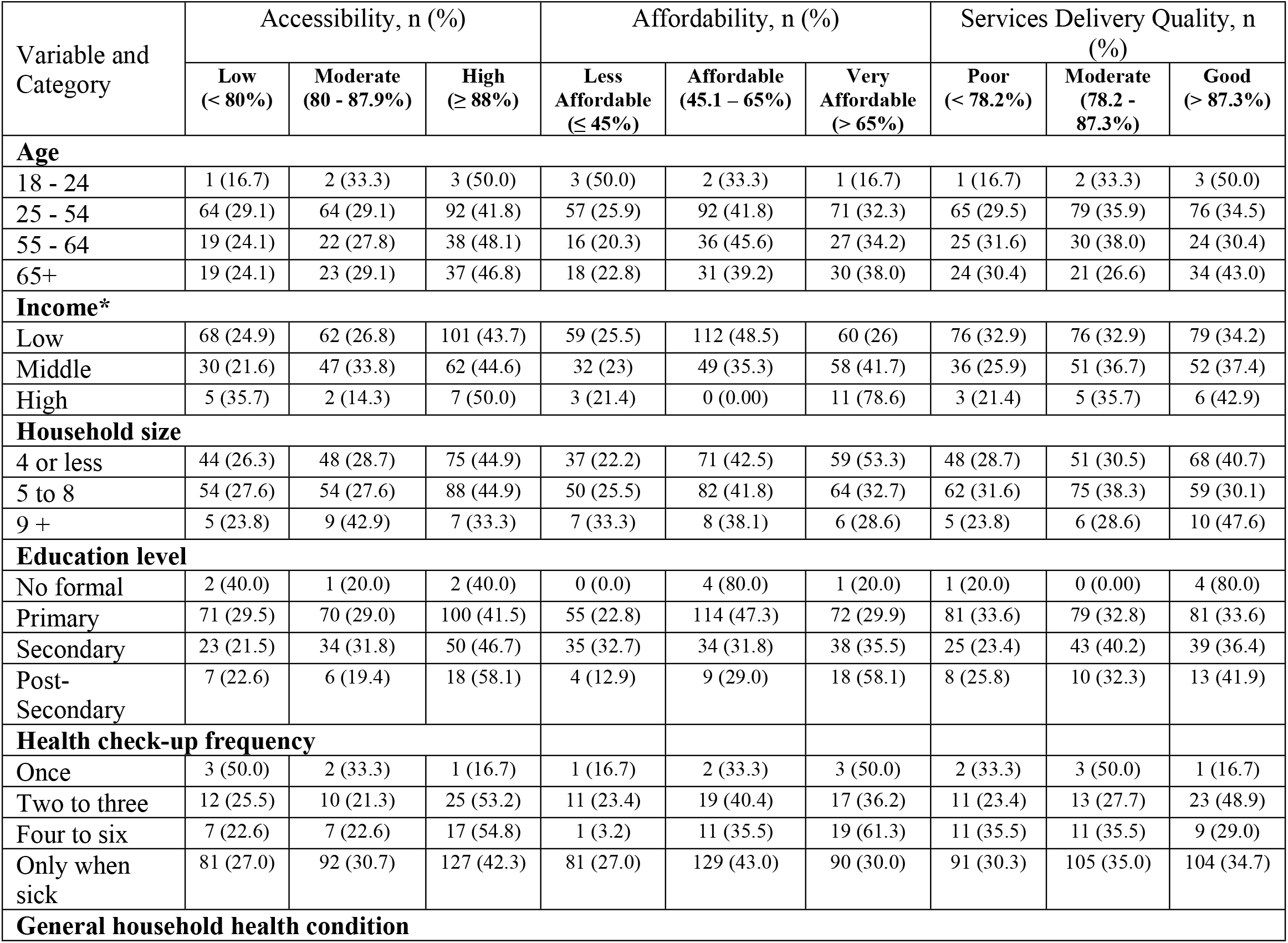
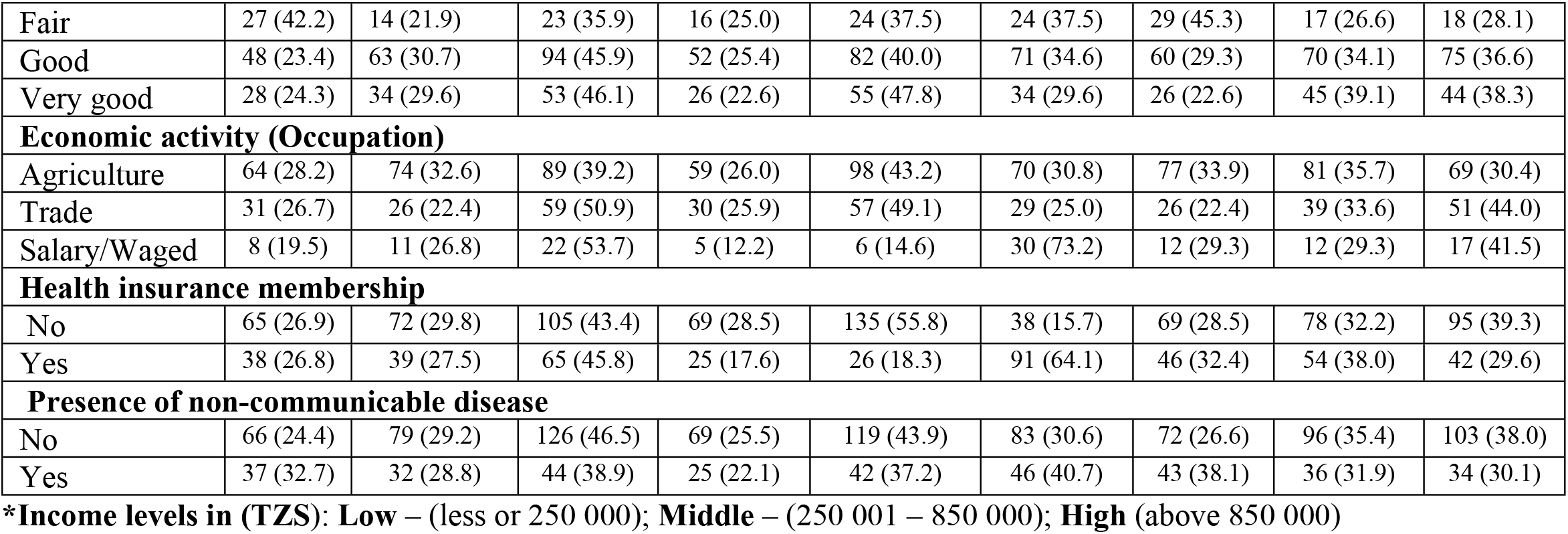
Socio-demographic characteristics and UHC factors crosstabulation (n = 384)

Majority of the household respondents under different socio-demographic characteristics as presented in Table 3 perceive the cost of healthcare services to be affordable with scores ranging from 45.1% to 65%. The respondents who perceived healthcare services to be very affordable (> 65%) fall in the socio-demographic categories of middle income 58 (41.7%) and high income 11 (78.6%) of the respondents. It is very possible that most of the households with low-income perceived health services to be affordable implying that they could only moderately afford the cost of healthcare services. Households with higher incomes were capable of securing the best services at any time compared with those with low income. The findings by [50] also observed that households with higher incomes were in the highest category of service affordability. Moreover, perceptions of healthcare services being very affordable were observed in the categories of household respondents’ education level (secondary and post-secondary). These findings resonate with findings of another study that education is related to once ability to afford healthcare cost [15]. Others are health check-up frequency (four to six times visits), occupation (salaried/waged), health insurance membership (yes category) and presence of non-communicable illness (yes category). The household respondents’ perceptions on service affordability (at very affordable) in relation with the socio-demographic features of income, education level, occupation, and health check-up frequency were also found to be significantly associated with health services affordability as presented in Table 4.

**Table 4:** Association between UHC factors and household socio-demographic features – Kruskal-Wallis H-test results (n = 384)

| Variable and Category | Accessibility |  | Affordability |  | Services delivery quality |  |
| --- | --- | --- | --- | --- | --- | --- |
|  | Mean Rank | H test (p -value) | Mean Rank | H test (p -value) | Mean Rank | H test (p -value) |
| Occupation |  |  |  |  |  |  |
| Agriculture | 183.27 | 4.210 (0.122) | 183.8 | 35.197 (0.000)*** | 179.31 | 8.843 (0.012)** |
| Trade | 203.28 |  | 175.74 |  | 216.69 |  |
| Salary/Waged | 213.07 |  | 288.12 |  | 197.12 |  |
| General household health condition |  |  |  |  |  |  |
| Fair | 161.08 | 6.368 (0.041)** | 199.14 | 0.693 (0.707) | 161.80 | 6.493 (0.039)** |
| Good | 198.44 |  | 194.13 |  | 195.12 |  |
| Very good | 199.40 |  | 185.90 |  | 204.92 |  |
| Education level |  |  |  |  |  |  |
| No Formal education | 169.30 | 1.735 (0.629) | 210.80 | 13.142 (0.004)*** | 235.00 | 4.786 (0.188) |
| Primary education | 188.45 |  | 184.86 |  | 183.37 |  |
| Secondary education | 197.06 |  | 189.26 |  | 208.21 |  |
| Post-Sec. education | 212.02 |  | 260.11 |  | 202.44 |  |
| Income |  |  |  |  |  |  |
| Low (less or 250 000) | 187.27 | 1.343 (0.511) | 177.19 | 17.345 (0.000)*** | 185.27 | 2.475 (0.290) |
| Middle (250 001 – 850 000) | 200.08 |  | 208.66 |  | 203.55 |  |
| High (Above 850 000) | 203.5 |  | 284.64 |  | 201.96 |  |
| Health check-up frequency |  |  |  |  |  |  |
| Once | 98.00 | 7.325 (0.062) | 219.83 | 16.227 (0.001)*** | 162.67 | 3.135 (0.371) |
| Two to three times | 205.90 |  | 196.66 |  | 206.73 |  |
| Four to six times | 219.48 |  | 265.97 |  | 165.21 |  |
| Only when I feel sick | 189.50 |  | 183.71 |  | 193.69 |  |

\*\*\* Association was significant at the 0.01 level (2-tailed)
\*\* Association was significant at the 0.05 level (2-tailed)

It can be noted from Table 3 that most of the household respondents’ perceptions on health services delivery quality were mixed between moderate (78.2% - 87.3%) and good (> 87.3%) among different categories of socio-demographic variables. While the majority of respondents with secondary education, for example, perceived health services delivery quality to be moderate, most of those with post-secondary education 13 (41.9%) perceived health services delivery quality to be good. Most of the households across all the health check-up categories perceived that health services delivery quality was moderate. The finding on health services delivery quality is in line with those of [51] who found that persons who went for health check-up more often perceived health services delivery quality to be moderately good. Moreover, while most of the respondents involved in agriculture perceived health services delivery quality as moderate, most of those in trade and salaried/waged occupations perceived health services delivery quality to be good. Whereas almost two-fifths 95 (39.3%) of those without health insurance perceived health services delivery quality as good, almost two-fifths 54 (38.0%) of those with health insurance perceived health services delivery quality as moderate. Most of the households with non-communicable illness 43 (38.1%), perceived health services delivery quality to be poor. This implies that people suffering from non-communicable illness were more likely to be frequent users of health facilities, thus, they had more occasions of interacting with healthcare personnel, thus, possible to experience weaknesses in health services provision.

### Differences in UHC levels by socio-demographic characteristics

Results are presented in Table 4 Table 5 for the Kruskal-Wallis H-test and Man-Whitney U-test, respectively. The associations between households’ socio-demographic characteristics and the factors for UHC attainment presented and discussed.

**Table 5:** Association between UHC factors and household socio-demographic features – Man Whitney U test results (n = 384)

| Variable and Category | Accessibility |  | Affordability |  | Service delivery quality |  |
| --- | --- | --- | --- | --- | --- | --- |
|  | Mean Rank | U test (p -value) | Mean Rank | U test (p -value) | Mean Rank | U test (p -value) |
| Health insurance membership |  |  |  |  |  |  |
| No | 190.82 | 17588.0 (0.694) | 154.37 | 26409.0 (0.000)*** | 201.42 | 15024.0 (0.039)** |
| Yes | 195.36 |  | 257.48 |  | 177.31 |  |
| Presence of non-communicable disease |  |  |  |  |  |  |
| No | 196.73 | 14166.5 (0.240) | 187.07 | 13840.0 (0.134) | 200.32 | 13192.0 (0.032)** |
| Yes | 182.37 |  | 205.52 |  | 173.74 |  |

\*\*\* Association was significant at the 0.01 level (2-tailed)
\*\* Association was significant at the 0.05 level (2-tailed)

#### Health services accessibility

As seen in Table 4, there was statistically significant difference in health services accessibility among households with perceived fair, good and very good general health condition [H (2) = 6.368, p = 0.041], with a mean rank accessibility score of 161.08 for fair, 198.44 for good, and 199.4 for very good household health condition. The Dunn’s post-hoc test with Bonferroni correction revealed significant differences between fair and good, p = 0.017, and between fair and very good household health condition, p = 0. 024. Household health condition was found to be associated with health services accessibility. The results imply that households with good or very good health condition had higher probability to access health services than those with a fair health condition. A related study revealed that persons with chronic illnesses (in this case, a fair health condition) were less likely to access health services due to high costs of regular health check-up and treatment [52].

#### Health services affordability

Results Table 4 indicates that there was a statistically significant difference in health services affordability between different categories of household’s monthly income [H (2) = 17.345, p = 0.000], with a mean rank affordability score of 177.19 for low income (≤ TZS. 250 000), 208.66 for middle income (TZS 250,001 – 850 000) and 284.64 for high income (above TZS 850 000). Dunn’s pairwise tests adjusted using the Bonferroni correction depicted significant differences between all three pairs of income categories: low and middle incomes, p = 0.008, low and high incomes, p = 0.000, and middle and high incomes, p = 0.014. The results imply that the pairs of lower income (Mean = 177, p = 0.008) and middle income (Mean = 208, p = 0.000) had a lower probability to afford health services than those with higher income. The finding echoes those of a study in India that household wealth (income being part of it) was one of the strong predictors to maternal healthcare affordability, thus, its utilization [17].

Moreover, Table 4 shows that health services affordability was statistically significantly different between different categories of occupation of the respondent households [H (2) = 35.197, p = 0.000], with a mean rank affordability score of 183.8 for agriculture, 175.74 for trade and 288.12 for salaried/waged occupation. A post-hoc test using Dunn’s test with Bonferroni correction indicated significant differences between trade and salaried/waged occupation, p = 0.000, and agriculture and salaried/waged, p = 0.000). Having a salaried/waged occupation was more associated with health services affordability than other occupation categories. The findings can be linked with those of [13] who found that occupation was associated with having health insurance, thus, more likelihood to afford health services.

There was statistically significant difference in health services affordability among households whose heads had different education levels [H (3) = 13.142, p = 0.004], with a mean rank service affordability score of 210.8 for no formal education, 184.84 for primary education, 189.26 for secondary education, and 260.11 for post-secondary education. Dunn’s post-hoc test with Bonferroni correction revealed statistically significant differences between pairs of primary and post-secondary education, p = 0.000, and between pairs of secondary and post-secondary education, p = 0.002. Concerning the household head education level, the results imply that the higher the household’s head level of education the higher the association with health services affordability at the household level. The studies [15] and [25] also found that education was strongly associated with financial coverage, which enhances health services affordability.

Furthermore, there was statistically significant difference in affordability score between different categories of health check-up frequency by respondents [H (3) = 16.227, p = 0.001], with a mean rank service affordability score of 219.83 for those who could go for health check-up once, 196.66 for two to three times, 265.97 for four to six times, and 183.71 for those who could go for health check-up only when they feel sick. A pairwise comparison using Dunn’s post-hoc test indicated significant differences between pairs of “only when I feel sick” and four to six times, p = 0.000, and between two to three times and four to six times, p = 0.006. The implication of the association between the household’s health check-up frequency and services affordability is that that household members who could go for health check-up at least once in a year were more likely to afford the cost of health services compared to those who could go for health check-up only when feeling sick. Similar studies indicate that frequent health check-up implies health services affordability by the health care users [42, 51].

Results Table 5 indicate that health services affordability was significantly higher for households with health insurance (Mean Rank = 257.48) than for households without health insurance (Mean Rank = 154.37), [U = 26409.0, p = 0.000]. It can be inferred from the results that participants with no health insurance were less likely to afford the cost of health services than those with health insurance. Other studies [16, 29] found that having medical insurance was associated with health seeking because it makes affordable for members to access healthcare more conveniently.

### Health services delivery quality

There was statistically significant difference in health services delivery quality among different occupation categories of the households Table 4 [H (2) = 8.843, p = 0.012], with a mean rank service delivery quality score of 179.31 for agriculture, 216.69 for trade and 197.12 for salaried/waged. From a pairwise comparison using Dunn’s post-hoc test with a Bonferroni correction, a combination involving agriculture and trade was significant, p = 0.003. However, there was no evidence of a difference between the other two pairs. The association between health services delivery quality and household’s head occupation implies that households involved in trade and salaried occupation were more likely to perceive good health services delivery quality because of their better financial statuses. Better financial status could enable them receive services of better quality.

Also, from Table 4 there was statistically significant difference in services delivery quality between different categories of general household health condition [H (2) = 6.493, p = 0.039], with a mean rank service delivery quality score of 161.8 for fair, 195.12 for good, and 204.92 for very good perception on services delivery quality among households. The Dunn’s post-hoc test with Bonferroni correction depicted significant differences between the pair of fair and good household health condition, p = 0.035, and that of fair and very good household health condition, p = 0.012. Households with very good health condition were more likely to rate higher the health services delivery quality than those with good or fair health condition. Perception of good health condition was associated with good health seeking behaviour [53].

Moreover, results Table 5 depict that perception on high health services delivery quality was statistically significantly associated with households with no health insurance (Mean Rank = 201.42) than for households with health insurance (Mean Rank = 177.31), [U = 15024.0, p = 0.039]. For the case of health insurance membership, there were occasions in some of the health facilities where clients who paid for services in cash were well served than those with health insurance, thus, perceived to have been served well. This underlines the observation in another study that use of health insurance in service access is associated with poor service provision by healthcare providers [54].

Furthermore, Table 5 shows that health services delivery quality was significantly higher for households without non-communicable disease (Mean Rank = 200.32) than for households with non-communicable disease (Mean Rank = 173.74), [U = 13192.0, p = 0.032]. Households without non-communicable diseases were more likely to perceive health service delivery quality as good because they may have been served well in their rare occasions of attending a health facility for healthcare. Not suffering from chronic illness implies less visits for health check-up [55], thus, less knowledgeable and experience about the shortcomings or strengths in health services delivery quality in health facilities.

### The level/extent of universal health coverage

Based on the geometric mean computation, the score for Reproductive, Martenal, Newborn and Child Health (RMNCH) domain was 94% and for Communicable Diseases Control (CDC) domain was 81.5% above the WHO’s recommended threshold of 80% score. The score for Non-Communicable Diseases Control (NCDC) domain was 57.1% and for Services Capacity and Access (SCA) domain was 54.7% below the WHO’s recommended threshold of 80% score. Therefore, considering the scores from the four domains, the UHC service coverage index for the study area was found to be 69.9%.

This score implies that the available health facilities in the four districts of Kilimanjaro Region had a coverage level of about 70% of health services provision. This score was highly contributed by the first two domains (Reproductive, Maternal, Newborn, and Child Health domain and Communicable Diseases Control domain) because it involves services provided in most of the health facilities, inlcuding in the health centres. The reproductive, maternal, newborn, and child health services are highly subsidized by the government in both government and private (especially faith-based) health facilities. For child immunization services, for example, the score was 100%, implying that the services were available at the required level in all the health facilities in the study area. This can be evidenced by the findings of a survey carried out in 2015/2016 in Tanzania indicating that Kilimanjaro Region was the leading region by 93% in Tanzania on provision of all basic vaccines to children aged 12-23 months [56]. The score, though measured based on selected health facilities in Kilimanjaro Region, was higher than the overall score of UHC service coverage index for Tanzania in 2017 which was 43% [57] compared to the minimum standard of 80% recommended by the WHO [58]. A study measuring effective UHC coverage in 124 countries in 2019 using 23 effective coverage indicators, indicated an improvement in the level of UHC coverage index for Tanzania to be 55% compared to that of 2017, which was 43% [5]. From the same study, it was found (similar to the current study) that Reproductive, Maternal, Newborn, and Child Health domain and Communicable Diseases Control domain performed better than the other domains [5].

## 4.0 Conclusions

Considering the study findings, majority of households in the study area could afford and access the needed health services at their convenience. Despite some negative perceptions on health services delivery quality by some of the household respondents, there was a general agreement that healthcare seekers could be served conveniently at the health facilities (health centres and hospitals) at any time. Health service providers should maintain the health services delivery quality as the local government authorities oversee health services accessibility and costs.

The significant households’ socio-demographic characteristics (education level, occupation, income, health insurance membership, presence of non-communicable illness, general household health condition and health check-up frequency) can predict the chances of a household to afford and access the needed health services. Improvement of the households’ social welfare is linked with health services accessibility and affordability as well as health services delivery quality. The Local Government Authorities (LGAs) should put in place enabling by-laws and conducive infrastructure for all people at the local level to access and utilize income generating opportunities. This would enable all people access the needed health services regardless of their socio-demographic characteristics.

Despite the fact that the score was highly contributed by the domain comprising reproductive, maternal, newborn, and child health as well as some components in communicable diseases control domain, the level of UHC in the study areas is fairly good. To improve progress towards attainment of UHC in the study area, the government, through the LGAs and Ministry of Health should stick to the set policy implementation. Mechanisms for improvement could be, among other, employing more and competent health workforce in the health facilities, enhance availability of modern medical equipment, improved health facilities infrastructure, and provision of universal health insurance to all household members.

## Data Availability

All data produced in the present study are available upon reasonable request to the authors.

## Authors’ contributions

Conceptualization: Kanti Ambrose Kimario, Kim A. Kayunze, and Mikidadi I. Muhanga.

Data organization, management, and analysis: Kanti Ambrose Kimario.

Investigation: Kanti Ambrose Kimario and Mikidadi I. Muhanga.

Methodology: Kanti Ambrose Kimario, Kim A. Kayunze, and Mikidadi I. Muhanga.

Project administration: Kanti Ambrose Kimario.

Resources: Kanti Ambrose Kimario.

Supervision: Kim A. Kayunze and Mikidadi I. Muhanga.

Validation: Kim A. Kayunze and Mikidadi I. Muhanga.

Writing – original draft: Kanti Ambrose Kimario, Kim A. Kayunze, and Mikidadi I. Muhanga.

Writing – review & editing: Kanti Ambrose Kimario, Mikidadi I. Muhanga, and Kim A. Kayunze.

**“S1 Table 1”: Proportionate random sampling of households. Legend:** CDH – Council Designated Hospital;

VAH – Voluntary Agency Hospital

**“S2 Table 2”: Health facilities selected from the four councils. Legend:** FBO – Faith-Based Organization; LGA – Local Government Authority; MC – Municipal Council; DC – District Council

**“S3 Table 3”: Measurement of independent variables**

**“S4 Table 4”: Measurement of dependent variable (UHC)**

**“S1 File 1”: Computation of UHC service coverage index based on geometric mean**

